# An exploratory intersectional analysis of syphilis prevalence among people who inject drugs in Montreal, Canada

**DOI:** 10.1101/2025.06.05.25328916

**Authors:** Brendan L. Harney, Julie Bruneau, Valérie Martel-Laferrière, Sarah Larney

**Author notes:** Corresponding author: Brendan Harney, Level 6, 900 Saint Denis St, Montreal, Quebec H2X 0A9.

## Abstract

**Background:** Syphilis notifications have increased among women and heterosexual men in Canada and people who inject drugs (PWID) are an emerging group at risk of infection. We examined syphilis prevalence among PWID and how this varied by intersecting population groups, socio-structural factors and sexual and substance use behaviors.

**Methods:** Data were from HEPCO, a cohort study of PWID in Montreal. We tested for syphilis via venipuncture with treponemal testing reflexed to non-treponemal testing if positive. We included the result at each person’s first test and exact logistic regression was used to examine differences in the prevalence of lifetime syphilis infection.

**Results:** Between November 2022 and March 2024, 386 people (16.1% women) had a syphilis test. Thirty-three people (8.6%) had lifetime syphilis and two people (0.5%) had active syphilis. Prevalence was significantly higher among men identifying as gay or bisexual, men with HIV, and those who reported recent sex work, condom-less sex, 2 or more sex partners and amphetamine injecting. Among women with syphilis, prevalence was higher among those who reported recent sex work, recent unstable housing and amphetamine injecting; however, the confidence interval included the null.

**Conclusion:** Active syphilis infection was uncommon among this cohort of PWID; however, lifetime syphilis infection was almost 9%. Among men in this cohort, there is evidence that syphilis intersects with other key population group characteristics. More data are needed to better understand syphilis among women who inject drugs. Periodic syphilis testing among PWID may be justified alongside testing for other sexually transmissible and blood borne infections.

**What is already known on this topic:** Contemporary increases in syphilis infection notifications in high income countries has been linked to substance use, including injecting drug use. However, there is a limited understanding of syphilis specifically among people who inject drugs.

**What this study adds:** This is the first study in more than 10 years to examine syphilis prevalence among people who inject drugs in Montreal, the second biggest city in Canada. Active syphilis was uncommon but still higher than the Canadian general population. Lifetime syphilis was associated with a range of intersecting key population groups and behaviours.

**How this study might affect research, practice or policy:** Our study provides a contemporary understanding of syphilis specifically among people who inject drugs in Montreal. Syphilis testing may be warranted among people who inject drugs to ensure prevalence stays low and guidelines from the Quebec government dating from 2019 should be revised.

## BACKGROUND

Once a disease that was thought to be possible to eliminate in Canada (1), syphilis rates have increased substantially in recent years; from 5.1 per 100,000 in 2011 to 24.7 per 100,000 in 2020 (2) and 36.1 per 100,000 in 2022 (3). While gay and bisexual men remain disproportionately represented in syphilis notifications in Canada, the most substantial increases have been among women (2). A consequence of this increase in syphilis among women has been a resurgence of congenital syphilis, from 0.3 to 31.7 cases per 100,000 births between 1993 and 2022(4). While this is likely driven by a range of factors at both an individual and system level, there is a growing body of evidence linking syphilis among women and heterosexual men to substance use, including injection drug use.

In Alberta, a province with one of the highest rates of syphilis in Canada, 46% of men who have sex with women and 44% of women diagnosed with syphilis in 2018 and 2019 reported lifetime stimulant use, compared to approximately 25% of gay and bisexual men (5). While stimulants can be consumed via numerous methods, stimulant use was associated with injection drug use and also having a sex partner who injects drugs. These findings align with data from the United States where there have been increases in the number of men who have sex with women and women diagnosed with syphilis reporting methamphetamine use and injecting drug use (6). In Australia, an analysis of data from 34 sexual health services between 2011 and 2019 found an association between injection drug use and syphilis among women (7). Based on these findings, it has been suggested that syphilis testing should be offered in services used by people who use drugs, including people who inject drugs. Despite this, there is a very limited contemporary understanding of syphilis specifically among people who inject drugs in Canada, and many other countries.

A systematic review and meta-analysis of syphilis and other STIs among people who inject drugs reported the global prevalence of active syphilis to be 3.2% (Price et al., 2025, Accepted, Drug and Alcohol Dependence). However, there was substantial variation across regions with the prevalence in North American studies being 0.1%. The most recent estimate of syphilis prevalence among people who inject drugs in Montreal dates from 2011/12 (8). Among 109 people, two had evidence of active syphilis infection, a prevalence of 1.8%.

In addition to a limited understanding of the contemporary epidemiology of syphilis among people who inject drugs, there is also a paucity of research exploring intersections between people who inject drugs and other key population groups at increased risk of syphilis, such as gay and bisexual men and sex workers. There also remains a lack of research exploring how social and structural determinants of health such as housing and incarceration may influence syphilis among people who inject drugs. We therefore aimed to examine the prevalence of syphilis among people who inject drugs in Montreal and explore how this differed according to intersecting population groups, social and structural determinants of health, as well as sexual and substance use behaviours.

## METHODS

### Data source

Data were from the Hepatitis C Cohort (HEPCO), a longitudinal cohort study of people who inject drugs in Montreal, Canada established in 2004 (9). People who are 18 years or older, have injected drugs in the last six months and are living in greater Montreal are eligible for enrolment. Recruitment is via street-based outreach, community partners and health care services. Participants are followed up every three months and each study visit included collection of venous blood samples and an interviewer administered questionnaire. HEPCO was approved by the research ethics board of the Centre hospitalier de l’Université de Montréal.

### Syphilis testing

Syphilis testing via venepuncture was added to the HEPCO study protocol in November 2022. Syphilis was detected using an anti-treponemal antibody test (Syphilis IgG EIA, Bio- Plex 2200 enzyme immunoassay, Bio-Rad), a rapid plasma reagin test (RPR carbon antigen, Pulse Scientific). New reactive cases were confirmed at the Laboratoire de Santé du Québec with a treponemal-specific test (TP-PA. +/- INNO-LIA). All reactive anti-treponemal antibody test were reviewed for the detection of active syphilis by a microbiologist infectious disease specialist, based on the longitudinal evolution of RPR titer, past medical and treatment history.

### Outcome – Lifetime syphilis infection

The primary outcome examined was lifetime syphilis infection inclusive of both active infection and historical infection, i.e., treponemal antibody positive. We included the syphilis result at each participants’ first study visit between November 2022 and March 2024.

### Exposure variables

We selected exposures and population groups for analyses based on published literature and syphilis testing guidelines (10). This included demographics (age, country of birth, Indigenous identity), social and structural determinants of health (recent incarceration and unstable housing), other infectious diseases (hepatitis C antibody and HIV status), other key population groups who may also be people who inject drugs (gay and bisexual men and sex workers) and sexual and substance use behaviours (recent condom use, number of sex partners and amphetamine injecting).

### Statistical analyses

Syphilis prevalence at the first test per participant and associated 95% confidence intervals were estimated using exact methods. Exact logistic regression was used to examine associations between lifetime syphilis infection for categorical variables and students t-test was used for continuous variables.

These analyses were stratified by gender which is worded in the questionnaire as “male or man” and “female or woman”. Other available response options are transgender, non-binary and two-spirit. Given the small number of people who did not identify as male/man or female/woman, we could not conduct a stratified analysis for this group. In a supplementary sensitivity analysis, we conducted the analyses based on self-reported sex assigned at birth.

### Missing data

For the outcome of lifetime syphilis infection, we compared the proportion missing across the exposure variables of interest to identify any potential bias that may be introduced. The proportion of missing data for the exposure variables was under 5% and as we were not undertaking multivariable analysis which could result in cumulative missing data for a participant (11), a complete case analysis was deemed acceptable.

All analyses were conducted using Stata18.5 SE (College Station, Texas, USA).

## RESULTS

Overall, 386 of 418 (92.3%) people had a valid syphilis test result available at their first study visit between November 2022 and March 2024. There were minimal differences in the proportion of people who did and did not have a valid syphilis test across the exposure variables of interest (Supplementary results table 1). Among the participants with a valid syphilis test, the mean age was 47 years (Std. Dev. 10.2, Range, 23-74) and the majority, 80.3%, identified as a male or man. Almost all were born in Canada and non-Indigenous and approximately half, 48.8%, reported recent unstable housing (Table 1).

**Table 1.** Overall characteristics of people who inject drugs tested for syphilis.

|  |  |
| --- | --- |
| Mean age (Std. Dev., Range) | 47 (10.2, 23-74) |
| Sex assigned at birth |  |
| Male | 322 (83.4) |
| Female | 64 (16.6) |
| Gender |  |
| Male or man | 310 (80.3) |
| Female or woman | 62 (16.1) |
| Non-binary/Transgender/two-spirit | 10 (2.6) |
| Missing | 4 (1.0) |
| Country of birth |  |
| Canada | 361 (93.5) |
| Other | 23 (6.0) |
| Missing | 2 (0.5) |
| Indigenous |  |
| No | 370 (95.9) |
| Yes | 16 (4.1) |
| Housing<sup>a</sup> |  |
| Stable | 198 (51.3) |
| Unstable | 188 (48.7) |
| Incarceration<sup>a</sup> |  |
| No | 346 (89.6) |
| Yes | 34 (8.8) |
| Missing | 6 (1.6) |
| Sexual identity |  |
| Heterosexual | 319 (82.6) |
| Gay or bisexual | 51 (13.2) |
| Missing | 16 (4.1) |
| HIV Status |  |
| Negative | 343 (88.9) |
| Positive | 39 (10.1) |
| Missing | 4 (1.0) |
| HCV antibody status |  |
| Negative | 153 (39.6) |
| Positive | 225 (58.3) |
| Missing | 8 (2.1) |
| Amphetamine injecting<sup>a</sup> |  |
| No | 277 (71.8) |
| Yes | 108 (28.0) |
| Missing | 1 (0.3) |
| Sex work<sup>a</sup> |  |
| No | 344 (89.1) |
| Yes | 40 (10.4) |
| Missing | 2 (0.5) |
| Condom use<sup>b</sup> |  |
| No sex | 241 (62.4) |
| Always | 39 (10.1) |
| Inconsistent | 106 (27.5) |
| Number of sexual partners<sup>b</sup> |  |
| No sex | 241 (62.4) |
| One | 79 (20.5) |
| Two or more | 63 (16.3) |
| Missing | 3 (0.8) |

Two people (0.5% [95%CI 0.1,2.1]) had an active syphilis infection and 33 people had evidence of lifetime syphilis infection, a prevalence of 8.6% (95%CI 6.1,11.8). The prevalence of lifetime syphilis infection was 8.1% among both men (95%CI 5.5,11.7) and women (95%CI 3.3,18.2), and 30% (95%CI 8.3,67.1) among non-binary, transgender or two-spirit participants.

Specific to participants who identified as men, the prevalence of lifetime syphilis infection was significantly higher among those who were living with HIV (Odds Ratio (OR) 6.7, 95% Confidence Internal [95%CI] 2.4,18.2)), identified as gay or bisexual (OR 8.0, 95%CI 2.9,22.0) and those who reported recent sex work (OR 8.7, 95%CI 2.6,27.7). It was also higher among those who reported amphetamine injecting (OR 2.9, 95%CI 1.2,7.3), inconsistent condom use (OR 5.6, 95%CI 2.1,15.5) and two or more sexual partners in the last month (OR 6.7, 95%CI 2.3,19.7) (Table 2).

**Table 2.**
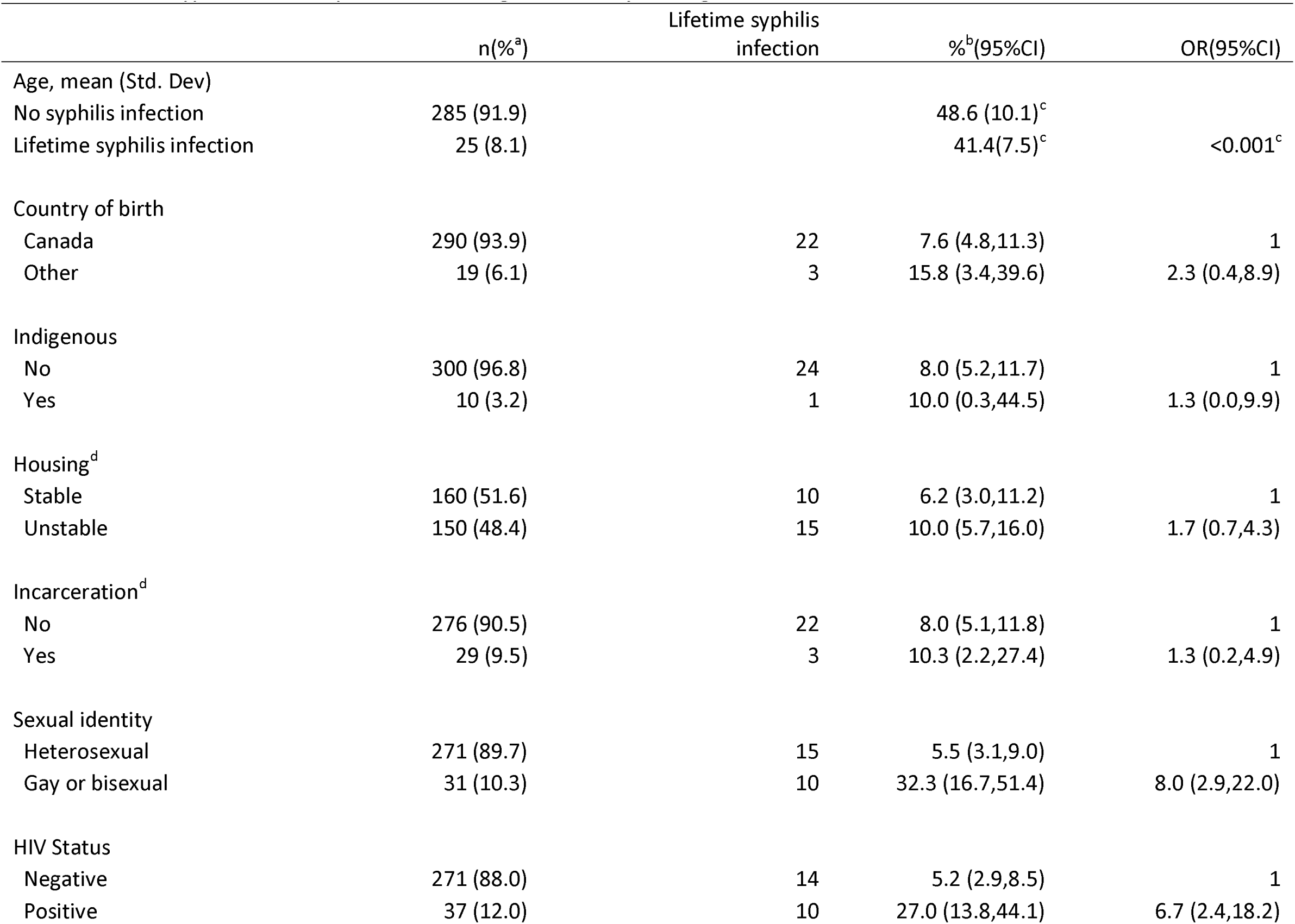
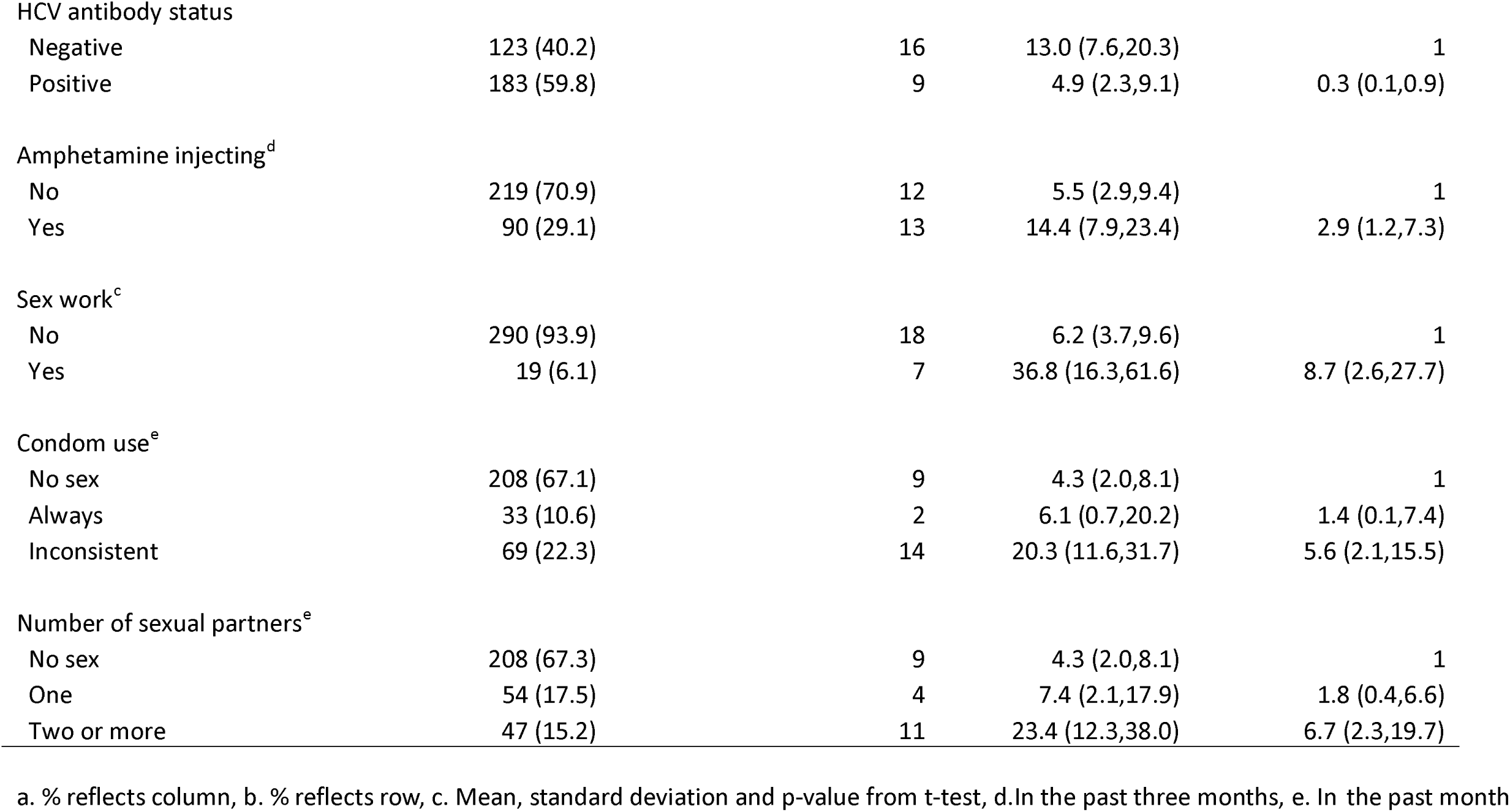
Lifetime syphilis infection prevalence among men who inject drugs in Montreal from November 2022 to March 2024, N=310.

|  | n(%<sup>a</sup>) | Lifetime syphilis<br>infection | %<sup>b</sup>(95%CI) | OR(95%CI) |
| --- | --- | --- | --- | --- |
| Age, mean (Std. Dev) |  |  |  |  |
| No syphilis infection | 285 (91.9) |  | 48.6 (10.1)<sup>c</sup> |  |
| Lifetime syphilis infection | 25 (8.1) |  | 41.4(7.5)<sup>c</sup> | <0.001<sup>c</sup> |
| Country of birth |  |  |  |  |
| Canada | 290 (93.9) | 22 | 7.6 (4.8,11.3) | 1 |
| Other | 19 (6.1) | 3 | 15.8 (3.4,39.6) | 2.3 (0.4,8.9) |
| Indigenous |  |  |  |  |
| No | 300 (96.8) | 24 | 8.0 (5.2,11.7) | 1 |
| Yes | 10 (3.2) | 1 | 10.0 (0.3,44.5) | 1.3 (0.0,9.9) |
| Housing<sup>d</sup> |  |  |  |  |
| Stable | 160 (51.6) | 10 | 6.2 (3.0,11.2) | 1 |
| Unstable | 150 (48.4) | 15 | 10.0 (5.7,16.0) | 1.7 (0.7,4.3) |
| Incarceration<sup>d</sup> |  |  |  |  |
| No | 276 (90.5) | 22 | 8.0 (5.1,11.8) | 1 |
| Yes | 29 (9.5) | 3 | 10.3 (2.2,27.4) | 1.3 (0.2,4.9) |
| Sexual identity |  |  |  |  |
| Heterosexual | 271 (89.7) | 15 | 5.5 (3.1,9.0) | 1 |
| Gay or bisexual | 31 (10.3) | 10 | 32.3 (16.7,51.4) | 8.0 (2.9,22.0) |
| HIV Status |  |  |  |  |
| Negative | 271 (88.0) | 14 | 5.2 (2.9,8.5) | 1 |
| Positive | 37 (12.0) | 10 | 27.0 (13.8,44.1) | 6.7 (2.4,18.2) |
| HCV antibody status |  |  |  |  |
| Negative | 123 (40.2) | 16 | 13.0 (7.6,20.3) | 1 |
| Positive | 183 (59.8) | 9 | 4.9 (2.3,9.1) | 0.3 (0.1,0.9) |
| Amphetamine injecting<sup>d</sup> |  |  |  |  |
| No | 219 (70.9) | 12 | 5.5 (2.9,9.4) | 1 |
| Yes | 90 (29.1) | 13 | 14.4 (7.9,23.4) | 2.9 (1.2,7.3) |
| Sex work<sup>c</sup> |  |  |  |  |
| No | 290 (93.9) | 18 | 6.2 (3.7,9.6) | 1 |
| Yes | 19 (6.1) | 7 | 36.8 (16.3,61.6) | 8.7 (2.6,27.7) |
| Condom use<sup>e</sup> |  |  |  |  |
| No sex | 208 (67.1) | 9 | 4.3 (2.0,8.1) | 1 |
| Always | 33 (10.6) | 2 | 6.1 (0.7,20.2) | 1.4 (0.1,7.4) |
| Inconsistent | 69 (22.3) | 14 | 20.3 (11.6,31.7) | 5.6 (2.1,15.5) |
| Number of sexual partners<sup>e</sup> |  |  |  |  |
| No sex | 208 (67.3) | 9 | 4.3 (2.0,8.1) | 1 |
| One | 54 (17.5) | 4 | 7.4 (2.1,17.9) | 1.8 (0.4,6.6) |
| Two or more | 47 (15.2) | 11 | 23.4 (12.3,38.0) | 6.7 (2.3,19.7) |

a. % reflects column, b. % reflects row, c. Mean, standard deviation and p-value from t-test, d. In the past three months, e. In the past month

Of the 25 men with a lifetime syphilis infection only 3 did not have one of the characteristics associated with increased prevalence, while of the other 22 men, 12 had three or more and six had two of these characteristics. Among the 14 transgender, non-binary or two-spirit participants, 12 reported male sex assigned at birth, and the three people with lifetime syphilis infection were also among this group. In the sensitivity analysis based on sex assigned at birth, the associations were the same as the primary analysis based on gender, however most were stronger (Supplementary table 2).

All participants who identified as women with lifetime syphilis infection were born in Canada, did not identify as Indigenous, were heterosexual and none were living with HIV. There was evidence that prevalence of lifetime syphilis infection was higher among those who reported recent unstable housing (10.3%), recent sex work (14.3%) and amphetamine injecting (15.4%); however, the confidence intervals were wide and included the null (Table 3). Of those with a lifetime syphilis infection, three had two or more of these characteristics, while two had none. As only two more people were included in the sensitivity analysis based on female sex assigned at birth, there was little difference in results (Supplementary table 3).

**Table 3.** Syphilis prevalence among women who inject drugs in Montreal from November 2022 to March 2024, N=62.

|  | n(%<sup>a</sup>) | Lifetime syphilis infection | %<sup>b</sup>(95%CI) | OR(95%CI) |
| --- | --- | --- | --- | --- |
| Age, mean (Std. Dev) |  |  |  |  |
| No syphilis infection | 57 (91.9) |  | 45.4 (10.0)<sup>c</sup> |  |
| Lifetime syphilis infection | 5 (8.1) |  | 47.8 (9.7)<sup>c</sup> | 0.60<sup>c</sup> |
| Country of birth |  |  |  |  |
| Canada | 58 (95.1) | 5 | 8.6 (2.9,19.0) | 1 |
| Other | 3 (4.9) | 0 | 0 (0.0,70.8) | 2.9 (0.0,31.1) |
| Indigenous |  |  |  |  |
| No | 58 (93.5) | 5 | 8.6 (2.9,19.0) | 1 |
| Yes | 4 (6.5) | 0 | 0 (0.0,60.2) | 2.2 (0.0,20.4) |
| Housing<sup>d</sup> |  |  |  |  |
| Stable | 33 (53.2) | 2 | 6.1 (0.7,20.2) | 1 |
| Unstable | 29 (46.8) | 3 | 10.3 (2.2,27.4) | 1.8 (0.2,22.7) |
| Incarceration<sup>d</sup> |  |  |  |  |
| No | 56 (91.8) | 4 | 7.1 (2.0,17.3) | 1 |
| Yes | 5 (8.2) | 1 | 20.0 (0.5,71.6) | 3.2 (0.1,45.4) |
| Sexual identity |  |  |  |  |
| Heterosexual | 43 (74.1) | 5 | 11.6 (3.9,25.1) | 1 |
| Gay or bisexual | 15 (25.9) | 0 | 0 (0.0,21.8) | 0.4 (0.0,3.1) |
| HIV Status |  |  |  |  |
| Negative | 59 (98.3) | 5 | 8.5 (2.8,18.7) | 1 |
| Positive | 1 (1.7) | 0 | 0 (0.0,97.5) | 11.0 (0.0,429.0) |
| HCV antibody status |  |  |  |  |
| Negative | 22 (37.3) | 1 | 4.5 (0.1,22.8) | 1 |
| Positive | 37 (62.7) | 4 | 10.8 (3.0,25.4) | 2.5 (0.2,131.2) |
| Amphetamine injecting<sup>d</sup> |  |  |  |  |
| No | 49 (79.0) | 3 | 6.1 (1.3,16.9) | 1 |
| Yes | 13 (21.0) | 2 | 15.4 (1.9,45.4) | 2.7 (0.2,27.0) |
| Sex work<sup>d</sup> |  |  |  |  |
| No | 47 (77.0) | 3 | 6.4 (1.3,17.5) | 1 |
| Yes | 14 (23.0) | 2 | 14.3 (1.8,42.8) | 2.4 (0.2,23.6) |
| Condom use<sup>e</sup> |  |  |  |  |
| No sex | 27 (43.5) | 2 | 7.4 (0.9,24.3) | 1 |
| Always | 5 (8.1) | 0 | 0 (0.0,52.2) | 2.2 (0.0,30.9) |
| Inconsistent | 30 (48.4) | 3 | 10.0 (2.1,26.5) | 1.4 (0.1,17.8) |
| Number of sexual partners<sup>e</sup> |  |  |  |  |
| No sex | 27 (44.3) | 2 | 7.4 (0.9,24.3) | 1 |
| One | 20 (32.8) | 2 | 10.0 (1.2,31.7) | 1.4 (0.1,20.7) |
| Two or more | 14 (23.0) | 1 | 7.1 (0.2,33.9) | 1.0 (0.0,20.2) |

1 a. % reflects column, b. % reflects row, c. Mean, standard deviation and p-value from t-test, d. In the past three months, e. In the past month

## DISCUSSION

In this cohort of 386 people who inject drugs, the prevalence of active syphilis was 0.5% and the prevalence of lifetime syphilis infection was 8.6%. The active syphilis prevalence of 0.5% was higher than that observed in a systematic review of STI prevalence among people who inject drugs with a 0.1% pooled prevalence in North American studies and similar to pooled results from studies in Western European countries of 0.6% (Price et al 2025, accepted, Drug and Alcohol Dependence). In comparison to the overall Canadian population, this is substantially higher than the national notification rate of 36.1 per 100,000 (0.036%) population and the Quebec specific rate of 14.1 per 100,000 (0.014%) (3).

We found evidence of intersections with other key populations and behaviours that would likely warrant syphilis testing regardless of injection drug use however there were some people not belonging to these groups or reporting these exposures who had lifetime syphilis infection. Among men, the prevalence of lifetime syphilis infection was significantly higher among those living with HIV and those who identified as gay or bisexual. Gay and bisexual men in general are over-represented in syphilis notification data in Canada (2). Of note, 32% of gay and bisexual men in our cohort had evidence of lifetime syphilis infection. This is substantially higher than the 18% prevalence reported among gay and bisexual men in Montreal from 2014-19 (12). Similarly, men living with HIV had a high prevalence in our cohort at 27%, approximately double the pooled prevalence of 13.2% from four Canadian studies of gay and bisexual men living with HIV (13).

Prevalence was also higher among men reporting recent inconsistent condom use, two or more sexual partners and amphetamine injecting. Though we cannot establish temporal proximity to previous infections, this suggests that there is an ongoing risk of new syphilis infections for these men. In addition, while the majority did not report any recent sex, of those who did, inconsistent condom use was common. Prevalence was also higher among males reporting sex work. While we cannot draw definitive conclusions, based on the strong overlap with gay and bisexual identity, this was mostly likely sex work involving other men (14).

While we were not able to conduct a stratified analysis for transgender, non-binary and two- spirit people, most of these participants reported male sex assigned at birth. Given the stronger associations generally found in our sensitivity analysis, it is likely that these would also be associated with an increased prevalence of syphilis among transgender, non-binary and two spirit people were the sample size large enough to conduct this analysis.

Among the women in our cohort, we found some evidence of differences in prevalence based on housing status, sex work and amphetamine use, however wide confidence intervals crossing the null prevent clear interpretation. A higher prevalence among women with unstable housing aligns with the increasing recognition of housing as social determinant of blood borne virus and sexually transmissible infection risk (15). Prevalence was also higher among women reporting recent sex work. While Canada does not criminalise sex workers per se, the criminalisation of clients of sex workers can often result in limited power for sex workers to negotiate sexual health practices such as condom use (16).

A higher prevalence of lifetime syphilis among women and men who reported injecting amphetamines (including methamphetamine) is notable. Historically amphetamine injecting has been limited among people who inject drugs in Montreal but annual prevalence increased from approximately 3% in 2011 to 13% in 2019 (17). Recent amphetamine injecting was reported by 28% of our sample, suggesting continued increases in recent years. Methamphetamine use has been associated with sexual behaviours that may in turn increase the risk of syphilis, and other STIs (18–20). Most of this evidence is however among gay and bisexual men, and there remains a limited understanding of this relationship among people who inject drugs generally, including heterosexual men and women. It is important to also recognise that some people, including women, may choose to use methamphetamine during sex for a variety of reasons including increasing desire and pleasure (19, 21). While our sample of women was small, and only five had lifetime syphilis infection, the overlap in exposures we found aligns with other evidence from this cohort showing a convergence of socio-structural risk factors and behaviours. For example, amphetamine use and sex work was more common among women who were unsheltered compared to those who were stably housed (22).

### Implications

National guidelines from the Public Health Agency of Canada published in 2024 suggest syphilis testing for “People who use substances and/or access addiction services”, which would clearly include people who inject drugs (10). There is however a note to “Consider local epidemiology when determining which groups/communities to target”. While evidence related to syphilis prevalence specifically among people who inject drugs is limited, some Canadian provincial guidelines do recommend testing among people who inject drugs (10).

Likewise, other countries seeing shifts in the epidemiology of syphilis such as Australia now also suggest that people who inject drugs should be tested (23).

Conversely, based on recommendations from 2019, injecting drug use is not an indication for syphilis testing in Quebec (24). However, it is important to note that syphilis has increased among the heterosexual population in recent years in Quebec (25). Furthermore, of 30 cases of congenital syphilis in Quebec between 2021 and 2023, 12 were among women who did not have syphilis screening early in pregnancy and/or were not treated during pregnancy (25). All of these women had a history of substance use, sex work, or homelessness which aligns closely with our findings. Strong links between congenital syphilis and birthing parent substance use have also been reported in other settings (26). In light of this, more nuanced and updated recommendations for syphilis testing among people who inject drugs in Quebec is likely warranted.

Given the limited attention bacterial STIs have received among people who inject drugs, there remains a lack of understanding of how best to provide care for syphilis, and other STIs, for this population. Evidence suggests that STI testing within needle and syringe programs is feasible (27) however more research is needed to understand this in the Canadian context and among a broader range of people who inject drugs. Given ongoing efforts to scale up hepatitis C testing and treatment, this could be an ideal opportunity to also introduce syphilis testing in this population. Rapid hepatitis C point of care testing has been shown to be highly acceptable and feasible among people who inject drugs in Canada (28) and similar high-income countries (29). Combining this with rapid testing for syphilis may have considerable benefits for people who inject drugs, their partners and their broader communities. This may be particularly pertinent given in some settings the overlap of syphilis and hepatitis C is substantial. For example, in Alberta, of 87 pregnant women who tested positive for hepatitis C, 17 (19.5%) also had an active syphilis infection (30). Similarly, the overlap in blood borne viruses and STIs is substantial in Manitoba where between 2018 and 2021 approximately 72% of females and 44% of males newly diagnosed with HIV reported injection drug use (31, 32). This further supports the suggestions and need to integrate harm reduction and sexual health services and make them accessible for people who inject drugs. Triangulation of data from such services, and bio-behavioural surveillance surveys, with the data from our cohort study will likely be of considerable value in further informing guidelines and service delivery related to syphilis among people who inject drugs.

## Limitations

Firstly, this cohort may not be representative of or generalisable to all people who inject drugs in Montreal. In particular, the mean age in our study was 47 and they may have stable partners and/or not be as sexually active as younger people who inject drugs. Second, there may be a degree of social desirability bias regarding sexual and/or substance use behaviours, particularly as this is an interviewer administrated questionnaire however some evidence suggests high concordance between biological testing and self-reported substance use behaviours (33). Thirdly, although our sample size was larger than other North American studies of syphilis among people who inject drugs, we were limited in our ability to conduct multivariable analyses, particularly among women, and have accordingly been conservative in our interpretation of findings.

In conclusion, these data provide a contemporary understanding of the prevalence of syphilis among a research engaged cohort of people who inject drugs in Montreal and provide a baseline for understanding a rapidly growing public health problem in Canada.

More data is needed to understand syphilis among women who inject drugs. Although the prevalence of active syphilis was low in this cohort, more person-centred testing in community settings may be of value to ensure this remains so given increasing rates in recent years.

## Supporting information

Strobe reporting guidelines

Supplementary results

## Data Availability

Data are not available outside of collaborative agreements due to Quebec privacy laws.

