## Supplementary results for "An exploratory intersectional analysis of syphilis prevalence among people who inject drugs in Montreal, Canada"

Supplementary table 1. Proportion of participants with and without a valid syphilis test result at their first study visit between November 2022 and March 2024

|  | Valid syphilis test % | No valid syphilis test % |
| --- | --- | --- |
| Sex assigned at birth |  |  |
| Male | 92.3 | 7.7 |
| Female | 92.8 | 7.2 |
| Gender identity |  |  |
| Male or man | 92.0 | 8.0 |
| Female or woman | 93.9 | 6.1 |
| Non-binary/Transgender/2-spirit | 90.9 | 9.1 |
| Country of birth |  |  |
| Canada | 92.6 | 7.4 |
| Other | 92.0 | 8.0 |
| Indigenous |  |  |
| No | 92.3 | 7.7 |
| Yes | 94.1 | 5.9 |
| Housing |  |  |
| Stable | 93.0 | 7.0 |
| Unstable | 91.7 | 8.3 |
| Incarceration |  |  |
| No | 92.5 | 7.5 |
| Yes | 97.1 | 2.9 |
| Sexual identity |  |  |
| Heterosexual | 93.0 | 7.0 |
| Gay or bisexual | 91.1 | 8.9 |
| HIV Status |  |  |
| Negative | 95.8 | 4.2 |
| Positive | 95.1 | 4.9 |
| HCV antibody status |  |  |
| Negative | 97.5 | 2.5 |
| Positive | 93.0 | 7.0 |
| Amphetamine injecting |  |  |
| No | 93.0 | 7.0 |
| Yes | 90.8 | 9.2 |
| Sex work |  |  |
| No | 94.0 | 6.0 |
| Yes | 83.3 | 16.7 |
| Condom use |  |  |
| No sex | 92.0 | 8.0 |
| Always | 92.9 | 7.1 |
| Inconsistent | 93.0 | 7.0 |
| Number of sexual partners | |  |
| No sex | 92.0 | 8.0 |
| One | 95.2 | 4.8 |
| Two or more | 90.0 | 10.0 |

Supplementary table 2. Lifetime syphilis infection prevalence among people assigned male sex at birth who inject drugs

in Montreal from November 2022 to March 2024, N=322

|  | n(%^a^) | Lifetime syphilis infection | %^b^(95%CI) | OR(95%CI) |
| --- | --- | --- | --- | --- |
| Country of birth |  |  |  |  |
| Canada | 302 (94.1) | 25 | 8.3 (5.4,12.0) | 1 |
| Other | 19 (5.9) | 3 | 15.8 (3.4,39.6) | 2.1 (0.4,8.0) |
| Indigenous |  |  |  |  |
| No | 309 (96.0) | 27 | 8.7 (5.8,12.5) | 1 |
| Yes | 13 (4.0) | 1 | 7.7 (0.2,36.0) | 0.9 (0.0,6.3) |
| Housing^c^ |  |  |  |  |
| Stable | 163 (50.6) | 11 | 6.7 (3.4,11.8) | 1 |
| Unstable | 159 (49.4) | 17 | 10.7 (6.4,16.6) | 1.7 (0.7,4.0) |
| Incarceration^c^ |  |  |  |  |
| No | 288 (90.9) | 25 | 8.7 (5.7,12.5) | 1 |
| Yes | 29 (9.1) | 3 | 10.3 (2.2,27.4) | 1.2 (0.2,4.4) |
| Sexual identity |  |  |  |  |
| Heterosexual | 277 (88.8) | 17 | 6.1 (3.6,9.6) | 1 |
| Gay or bisexual | 35 (11.2) | 10 | 28.6 (14.6,46.3) | 6.1 (2.2,15.9) |
| HIV status |  |  |  |  |
| Negative | 283 (88.4) | 17 | 6.0 (3.5,9.4) | 1 |
| Positive | 37 (11.6) | 10 | 27.0 (13.8,44.1) | 5.7 (2.1,14.9) |
| HCV antibody status |  |  |  |  |
| Negative | 130 (40.9) | 18 | 13.8 (8.4,21.0) | 1 |
| Positive | 188 (59.1) | 10 | 5.3 (2.6,9.6) | 0.4 (0.1,0.8) |
| Amphetamine injecting^c^ |  |  |  |  |
| No | 226 (70.4) | 12 | 5.3 (2.8,9.1) | 1 |
| Yes | 95 (29.6) | 16 | 16.8 (9.9,25.9) | 3.6 (1.5,8.7) |
| Sex work^c^ |  |  |  |  |
| No | 297 (92.5) | 18 | 6.1 (3.6,9.4) | 1 |
| Yes | 24 (7.5) | 10 | 41.7 (22.1,63.4) | 10.9 (3.8,31.0) |
| Condom use^d^ |  |  |  |  |
| No sex | 213 (66.1) | 10 | 4.7 (2.3,8.5) | 1 |
| Always | 33 (10.2) | 2 | 6.1 (0.7,20.2) | 1.3 (0.1,6.6) |
| Inconsistent | 76 (23.6) | 16 | 21.1 (12.5,31.9) | 5.4 (2.2,14.0) |
| Number of sexual partners^d^ |  |  |  |  |
| No sex | 213 (66.4) | 10 | 4.7 (2.3,8.5) | 1 |
| One | 58 (18.1) | 5 | 8.6 (2.9,19.0) | 1.9 (0.5,6.5) |
| Two or more | 50 (15.6) | 12 | 24.0 (13.1,38.2) | 6.3 (2.3,17.7) |

a. % reflects column, b. % reflects row, c. In the past three months, d. In the past month

Supplementary table 3. Lifetime syphilis infection prevalence among people assigned female sex at birth who inject drugs

in Montreal from November 2022 to March 2024, N=64

|  | n(%^a^) |  | Lifetime syphilis infection | %^b^(95%CI) | OR(95%CI) |
| --- | --- | --- | --- | --- | --- |
| Country of birth |  |  |  |  |  |
| Canada | 59 (93.7) |  | 5 | 8.5 (2.8,18.7) | 1 |
| Other | 4 (6.3) |  | 0 | 0 (0.0 60.2) | 2.2 (0.0,20.8) |
| Indigenous |  |  |  |  |  |
| No | 61 (95.3) |  | 5 | 8.2 (2.7,18.1) | 1 |
| Yes | 3 (4.7) |  | 0 | 0.0 70.8) | 3.1 (0.0,32.8) |
| Housing^c^ |  |  |  |  |  |
| Stable | 35 (54.7) |  | 2 | 5.7 (0.7,19.2) | 1 |
| Unstable | 29 (45.3) |  | 3 | 10.3 (2.2,27.4) | 1.9 (0.2,24.1) |
| Incarceration^c^ |  |  |  |  |  |
| No | 58 (92.1) |  | 4 | 6.9 (1.9,16.7) | 1 |
| Yes | 5 (7.9) |  | 1 | 20.0 (0.5,71.6) | 3.3 (0.1,47.1) |
| Sexual identity |  |  |  |  |  |
| Heterosexual | 42 (72.4) |  | 5 | 11.9 (4.0,25.6) | 1 |
| Gay or bisexual | 16 (27.6) |  | 0 | 0 (0.0 20.6) | 0.4 (0.0,2.8) |
| HIV status |  |  |  |  |  |
| Negative | 60 (96.8) |  | 5 | 8.3 (2.8,18.4) | 1 |
| Positive | 2 (3.2) |  | 0 | 0 (0.0 84.2) | 4.8 (0.0,65.9) |
| HCV antibody status |  |  |  |  |  |
| Negative | 23 (38.3) |  | 1 | 4.3 (0.1,21.9) | 1 |
| Positive | 37 (61.7) |  | 4 | 10.8 (3.0,25.4) | 2.6 (0.2,137.2) |
| Amphetamine injecting^c^ | |  |  |  |  |
| No | 51 (79.7) |  | 3 | 5.9 (1.2,16.2) | 1 |
| Yes | 13 (20.3) |  | 2 | 15.4 (1.9,45.4) | 2.8 (0.2,28.2) |
| Sex work^c^ |  |  |  |  |  |
| No | 47 (74.6) |  | 3 | 6.4 (1.3,17.5) | 1 |
| Yes | 16 (25.4) |  | 2 | 12.5 (1.6,38.3) | 2.1 (0.2,20.0) |
| Condom use^d^ |  |  |  |  |  |
| No sex | 28 (43.8) |  | 2 | 7.1 (0.9,23.5) | 1 |
| Always | 6 (9.4) |  |  | 0 (0.0 45.9) | 1.9 (0.0,26.2) |
| Inconsistent | 30 (46.9) |  | 3 | 10.0 (2.1,26.5) | 1.4 (0.2,18.5) |
| Number of sexual partners^d^ | |  |  |  |  |
| No sex | 28 (45.2) |  | 2 | 7.1 (0.9,23.5) | 1 |
| One | 21 (33.9) |  | 2 | 9.5 (1.2,30.4) | 1.4 (0.1,20.3) |
| Two or more | 13 (21.0) |  | 1 | 7.7 (0.2,36.0) | 1.1 (0.0,22.7) |

a. % reflects column, b. % reflects row, c. In the past three months, d. In the past month
